# Understanding of and Trust in the Centers for Disease Control and Prevention’s Revised COVID-19 Isolation and Quarantine Guidance Among US Adults

**DOI:** 10.1101/2022.02.01.22270288

**Authors:** Vishala Mishra, Joseph P. Dexter

**Affiliations:** Department of Biostatistics, Harvard T.H. Chan School of Public Health, Boston, Massachusetts; Data Science Initiative, Harvard University, Allston, Massachusetts

## Abstract

On December 27, 2021 the Centers for Disease Control and Prevention (CDC) announced changes to their guidance for individuals who are exposed to or test positive for COVID-19. The revised recommendations have prompted widespread discussion of both the scientific rationale and communication strategy, including criticism from the American Medical Association.^1,2^ In this survey study, we assessed understanding of and trust in the CDC’s initial public statement about the new guidance among US adults.

## Methods

We administered the online survey to 603 participants recruited through Prolific between January 5-6, 2022. The survey instrument is provided in eAppendix 1.

The cohort was assembled using nonprobability convenience sampling of US adults, with quotas chosen to match 2019 US Census data on age, race, ethnicity, and education (eMethods). The study was approved by Harvard University’s Committee on the Use of Human Subjects, and participants provided informed consent electronically before beginning the survey.

Associations between participant characteristics and passage comprehension were determined using ordinal logistic regression.

## Results

The demographic characteristics of the participants are listed in the Table. Participants answered comprehension questions about application of the isolation and quarantine guidelines to hypothetical scenarios, based on either a vaccination history specified in the question (“scenario” questions) or their own history (“personal” questions). 150 (25%) participants correctly answered all 4 scenario questions, and 180 (30%) participants correctly answered all 4 personal questions (Table). Being unvaccinated for COVID-19 was negatively associated with number of correct responses to both the scenario (odds ratio [OR], 0.74; 95% CI, 0.59-0.93; *P* = .01) and personal (OR, 0.61; 95% CI, 0.48-0.77; *P* < .001) questions; not having received a booster was negatively associated with number of correct responses to the personal questions (OR, 0.72; 95% CI, 0.58-0.88; *P* = .001).

**Table.**
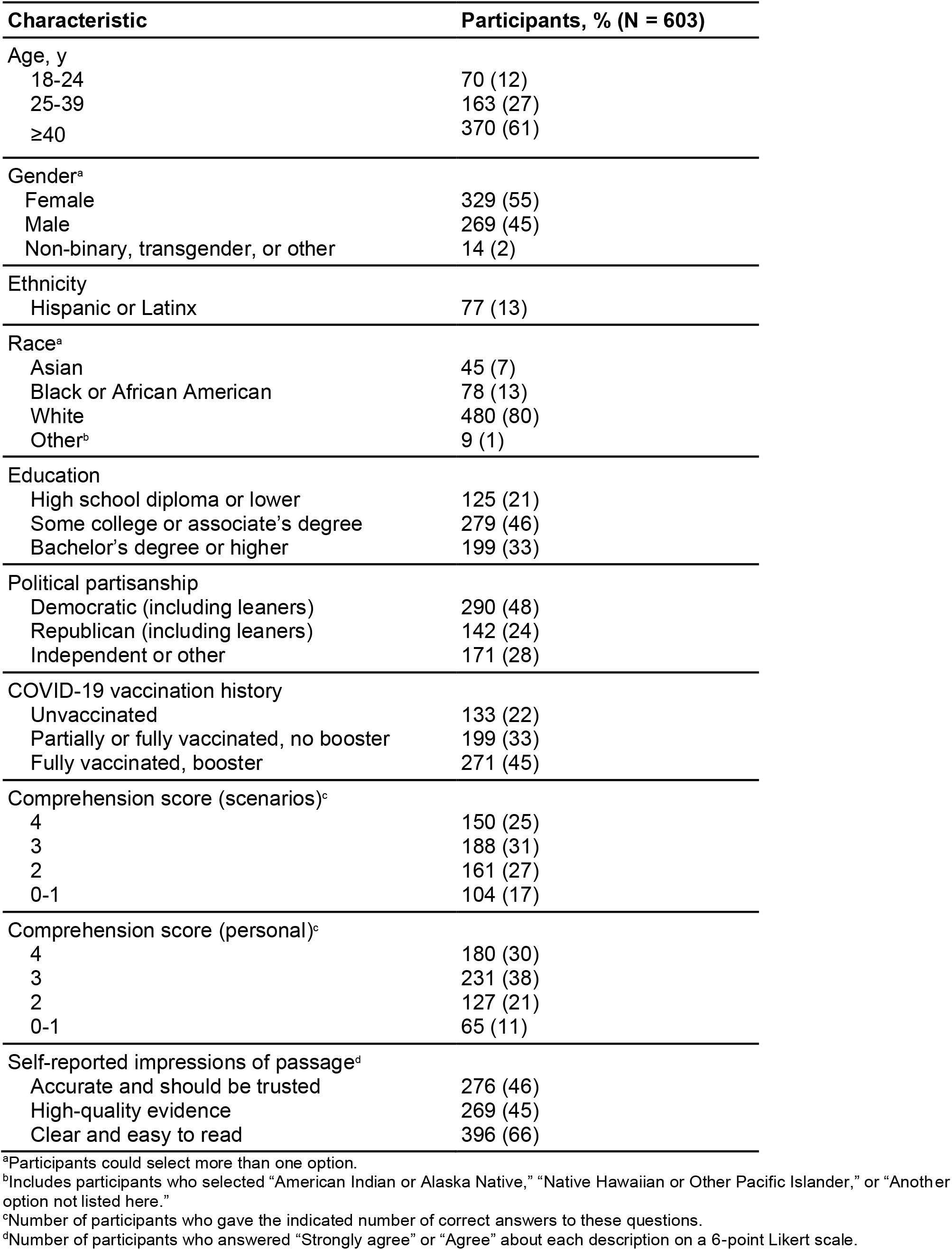
Characteristics and Responses of Participants Who Completed the Online Survey About Change in CDC Isolation and Quarantine Guidance.

The CDC web page stated that vaccination “decreases the risk of severe disease, hospitalization, and death from COVID-19” but gave quantitative estimates only for effectiveness against infection. When participants were asked to guess the effectiveness against hospitalization from COVID-19, the modal response was 30-39% without a booster (139 [23%] participants; Figure) and 70-79% with a booster (177 [29%] participants; Figure), corresponding to the stated numbers for effectiveness against infection (35% and 75%, respectively). A majority of participants guessed that vaccination is less than 90% effective against death from COVID-19 both without (437 [72%] participants) and with a booster (342 [57%]; Figure).

**Figure.**
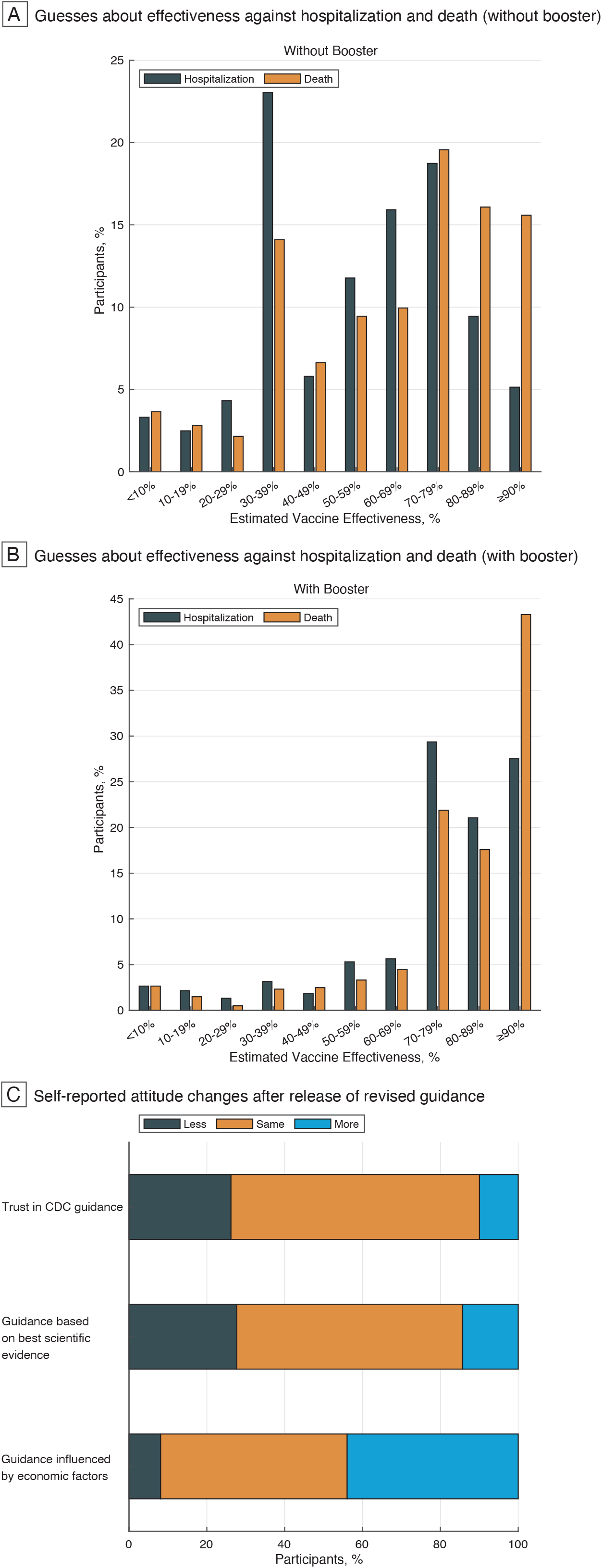
Self-Reported Attitude Changes and Estimated Vaccine Effectiveness Against Hospitalization or Death from COVID-19. The graphs show (A) estimated effectiveness of a COVID-19 vaccine without a booster against hospitalization (orange bars) or death (gray bars), (B) estimated effectiveness of a COVID-19 vaccine with a booster against hospitalization (orange bars) or death (gray bars), and (C) the percentage of respondents who expressed changes in attitude before and after release of the revised guidance in response to three counterfactual questions.

Participants were asked about their current attitudes towards the CDC’s COVID-19 guidance, as well as what their attitudes had been before announcement of the changes. In response to these counterfactual questions (eMethods),^3^ 158 (26%) participants indicated that the change in guidance lowered their overall trust of the CDC’s recommendations (Figure). 167 (28%) participants expressed reduced confidence that the agency relies on the best scientific evidence, and 265 (44%) said they now think it is more likely that economic factors influence CDC guidance (Figure).

## Discussion

Public health messaging about the Omicron variant must balance speed, clarity, and responsiveness to rapid scientific changes.^1,4^ In a survey of a representative sample of US adults, comprehension testing of the CDC’s revised guidance revealed widespread gaps in understanding. The negative association of comprehension scores with vaccination status suggests the recommendations may be least accessible to individuals at greatest risk of infection.

When asked counterfactual questions, many participants expressed reduced trust in CDC recommendations about COVID-19 and a stronger belief that the agency’s guidance is influenced by economic considerations. Participants also underestimated the protectiveness of COVID-19 vaccines against hospitalization and death, suggesting that specific numbers should have been provided on the web page to reduce variability in risk perception.^5^ This omission is notable in light of evidence that highlighting protection against death may reduce COVID-19 vaccine hesitancy.^6^

Limitations of the study include that it was conducted only online, such that individuals without internet access may have been undersampled, and use of a nonprobability sample, which limits generalizability to the US population as a whole.

## Supporting information

Supplemental Online Content

## Data Availability

Raw data is available from the corresponding author on request.

## Author Contributions

Drs Mishra and Dexter had full access to all of the data in the study and take responsibility for the integrity of the data and the accuracy of the data analysis.

*Concept and design:* Both authors.

*Acquisition, analysis, or interpretation of data:* Both authors.

*Drafting of the manuscript:* Both authors.

*Critical revision of the manuscript for important intellectual content:* Both authors.

*Statistical analysis:* Both authors.

*Obtained funding:* Dexter.

*Supervision:* Both authors.

## Conflict of Interest Disclosures

Dr Dexter reported receiving grants from the Poynter Institute and the Harvard Data Science Initiative during the conduct of the study. No other disclosures were reported.

## Funding/Support

This work was supported by a CoronaVirusFacts Alliance Grant from the Poynter Institute and a Harvard Data Science Fellowship.

## Role of the Funder/Sponsor

The funders had no role in the design and conduct of the study; collection, management, analysis, and interpretation of the data; preparation, review, or approval of the manuscript; and decision to submit the manuscript for publication.

## Notes

### Competing Interest Statement

The authors have declared no competing interest.

### Author Declarations

The study was approved by Harvard University's Committee on the Use of Human Subjects.

