## Supplemental Online Content for "Understanding of and Trust in the Centers for Disease Control and Prevention’s Revised COVID-19 Isolation and Quarantine Guidance Among US Adults"

**eMethods**

**eAppendix 1.** Survey Administered to Participants

This supplemental material has been provided by the authors to give readers additional information about their work.

**eMethods**

*CDC Passage*

The passage used in the survey was a press release posted online by the Centers for Disease Control and Prevention (CDC) on December 27, 2021. The full text of the passage is provided in eAppendix 1 and is also available at: https://www.cdc.gov/media/releases/2021/s1227-isolation-quarantine-guidance.html.

*Demographic Information*

We collected self-reported demographic information about age, gender, ethnicity, race, education, political partisanship, and COVID-19 vaccination history. Participants were asked about their race and ethnicity because of disparities in COVID-19 vaccine uptake across racial and ethnic minority groups.[1,2,3]

*Quota Sampling Strategy*

As noted in the main text, the cohort was assembled using nonprobability convenience sampling of US adults, with quotas chosen to match 2019 US Census data on age ("18-24," "25-39," "40+"), race ("Not White Alone" or "White Alone"), ethnicity ("Non-Hispanic" or "Hispanic"), and education ("No Bachelor's Degree" or "Bachelor's Degree").[4] On the basis of these divisions, we defined 24 non-overlapping group and set the size of each group to be proportional to the general population; group sizes were rounded to ensure that there would be at least 1 participant in every group.

For the sample of 603 participants, the group sizes were as follows:

Group 1 (18-24, White Alone, Non-Hispanic, No Bachelor's Degree): 32
Group 2 (18-24, White Alone, Hispanic, No Bachelor's Degree): 13
Group 3 (18-24, Not White Alone, Non-Hispanic, No Bachelor's Degree): 14
Group 4 (18-24, Not White Alone, Hispanic, No Bachelor's Degree): 2
Group 5 (25-39, White Alone, Non-Hispanic, No Bachelor's Degree): 47
Group 6 (25-39, White Alone, Hispanic, No Bachelor's Degree): 23
Group 7 (25-39, Not White Alone, Non-Hispanic, No Bachelor's Degree): 22
Group 8 (25-39, Not White Alone, Hispanic, No Bachelor's Degree): 3
Group 9 (40+, White Alone, Non-Hispanic, No Bachelor's Degree): 158
Group 10 (40+, White Alone, Hispanic, No Bachelor's Degree): 37
Group 11 (40+, Not White Alone, Non-Hispanic, No Bachelor's Degree): 46
Group 12 (40+, Not White Alone, Hispanic, No Bachelor's Degree): 4
Group 13 (18-24, White Alone, Non-Hispanic, Bachelor's Degree): 6
Group 14 (18-24, White Alone, Hispanic, Bachelor's Degree): 1
Group 15 (18-24, Not White Alone, Non-Hispanic, Bachelor's Degree): 2
Group 16 (18-24, Not White Alone, Hispanic, Bachelor's Degree): 1
Group 17 (25-39, White Alone, Non-Hispanic, Bachelor's Degree): 42
Group 18 (25-39, White Alone, Hispanic, Bachelor's Degree): 6

Group 19 (25-39, Not White Alone, Non-Hispanic, Bachelor’s Degree): 16
Group 20 (25-39, Not White Alone, Hispanic, Bachelor’s Degree): 1
Group 21 (40+, White Alone, Non-Hispanic, Bachelor’s Degree): 95
Group 22 (40+, White Alone, Hispanic, Bachelor’s Degree): 8
Group 23 (40+, Not White Alone, Non-Hispanic, Bachelor’s Degree): 23
Group 24 (40+, Not White Alone, Hispanic, Bachelor’s Degree): 1

Participants were recruited through Prolific (https://www.prolific.co/). Prescreening questions written by Prolific and previously administered to their participant base were used to limit enrollment to the target populations.

The following prescreening question was asked of all participants:

Question: “In which country do you currently reside?”

Accepted answer: “United States”

The following prescreening questions were used to determine race, ethnicity, and education for group eligibility:

Question: “What ethnic group do you belong to?”

Accepted answers (“Not White Alone”): “Black,” “Asian,” or “Mixed”

Accepted answer (“White Alone”): “White”

Question: “Please indicate your ethnicity (i.e. peoples’ ethnicity describes their feeling of belonging and attachment to a distinct group of a larger population that shares their ancestry, colour, language or religion)?”

Accepted answers (“Non-Hispanic”): “African,” “Black/African American,” “Caribbean,” “East Asian,” “Middle Eastern,” “Native American or Alaskan Native,” “South Asian,” “White/Caucasian,” “White/Sephardic Jew,” “Black/British,” “White Mexican,” “Romani/Traveller,” or “South East Asian”

Accepted answer (“Hispanic”): “Latino/Hispanic”

Question: Which of these is the highest level of education you have completed?

Accepted answers (“No Bachelor’s Degree”): “No formal qualifications,” “Secondary education (e.g. GED/GCSE),” “High school diploma/A-levels,” or “Technical/community college”

Accepted answers (“Bachelor’s Degree”): “Undergraduate degree (BA/BSc/other),” “Graduate degree (MA/MSc/MPhil/other),” or “Doctorate degree (PhD/other)”

All but 1 of the groups filled within 24 hours of the launch of the study; 9 participants enrolled in Group 10, which had a quota of 37. To compensate for this under-enrollment, we then recruited an additional 28 participants who were 40 years of age or older and had not received a bachelor's degree. This final group was restricted by education because of previous research indicating that health literacy varies more by educational attainment than by ethnicity.[5]

*Participant Compensation*

Participants received $2.00 after completion of the survey.

*Comprehension Questions*

We developed a series of multiple-choice questions to assess how well participants understood the isolation and quarantine recommendations in the CDC passage. For the first 4 questions, participants answered based on a vaccination history specified in the question ("scenario" questions); for the last 4 questions, they answered based on their own history ("personal" questions). The questions, which are provided in eAppendix 1, are of uniform format and involve identifying the correct behavior (isolation, quarantine, wearing a mask in public, or no changes to daily routine) in various hypothetical scenarios.

*Impression Questions*

Participants were asked to rate their impressions of various attributes of the passage using a 6-point Likert scale (options "Strongly disagree," "Disagree," "Slightly disagree," "Slightly agree," "Agree," and "Strongly agree"). We adapted these questions, which are provided in eAppendix 1, from a prior study of COVID-19 health literacy by Kerr *et al*. 2021.[6]

*Guess Questions*

The CDC announcement stated, "Data from South Africa and the United Kingdom demonstrate that vaccine effectiveness against infection for two doses of an mRNA vaccine is approximately 35%. A COVID-19 vaccine booster dose restores vaccine effectiveness against infection to 75%." The passage did not, however, give quantitative estimates of vaccine effectiveness against hospitalization or death from COVID-19. We asked participants to provide their best guesses for effectiveness against hospitalization and death (both with and without a booster dose).

*Counterfactual Questions*

We used the nonrandomized counterfactual format described in Graham and Coppock 2021 to measure participants' beliefs about the effect of the revised recommendations on their trust in the CDC's guidance about COVID-19, their confidence that the CDC relies on the best scientific evidence, and the extent to which CDC guidance is influenced by economic factors.[7] Participants first answered each question based on their current beliefs after reading the passage. They were then instructed to answer the same questions as if they had been surveyed before the revised recommendations were released.[7] For the figure, we determined the number of participants who expressed a stronger, weaker, or the same belief in response to the counterfactual questions than the baseline questions.

*Statistical analysis*

Associations between participant characteristics and performance on the scenario and personal comprehension questions were assessed using ordinal logistic regression. For the 2 models, the dependent variable was the total number of correct responses to the scenario and personal comprehension questions, respectively. The following features were included in each model (all categories correspond to those in the Table in the main text): “Age: ≥40,” “Gender: Female,” “Ethnicity: Hispanic of Latinx,” “Race: Asian,” “Race: Black or African American,” “Race: Other,” “Education: High school diploma or lower; Some college or associate’s degree,” “Political partisanship: Republican (including leaners),” “COVID-19 vaccination history: Unvaccinated,” and “COVID-19 vaccination history: Partially or fully vaccinated, no booster.”

For the scenario comprehension questions, the significant features were “Gender: Female” (odds ratio [OR], 1.21; 95% CI, 1.02-1.45; $P = .031$) and “COVID-19 vaccination history: Unvaccinated” (OR, 0.74; 95% CI, 0.59-0.93; $P = .01$). For the personal comprehension questions, the significant features were “Race: Asian” (OR, 1.46; 95% CI, 1.03-2.07; $P = .033$), “Political partisanship: Republican (including leaners)” (OR, 0.75, 95% CI, 0.48-0.77; $P = .009$), “COVID-19 vaccination history: Unvaccinated” (OR, 0.61; 95% CI, 0.48-0.77; $P < .001$), and “COVID-19 vaccination history: Partially or fully vaccinated, no booster” (OR, 0.72; 95% CI, 0.58-0.88; $P = .001$).

Statistical analyses were performed using the Python package statsmodels (version 0.14.0.dev0). Statistical significance was defined as $P < .05$.

**eAppdenix 1. Survey Administered to Participants**

In what country do you currently reside?
▼ United Kingdom ... Zimbabwe

*Please answer the following questions about your experiences with the COVID-19 pandemic and your vaccination history.*

When was the last time you visited the CDC's website?
- Within the past three days
- Within the past week
- Within the past two weeks
- More than two weeks ago
- I have never visited the CDC's website.

Have you gotten at least one shot of any COVID-19 vaccine?
- No
- Yes

Are you fully vaccinated for COVID-19? Fully vaccinated means getting <u>EITHER</u> *two* shots of the Pfizer-BioNTech or Moderna vaccine <u>OR</u> *one* shot of the J&J-Janssen vaccine. [Display Logic: If “Have you gotten at least one shot of any COVID-19 vaccine? = Yes”]
- No
- Yes

Which vaccine did you get for your primary (<u>NOT</u> booster) series? [Display Logic: If “Are you fully vaccinated for COVID-19? Fully vaccinated means getting EITHER two shots of the Pfi... = Yes”]
- Pfizer-BioNTech
- Moderna
- J&J-Janssen

When did you get the <u>LAST</u> shot of your primary (<u>NOT</u> booster) series? [Display Logic: If “Are you fully vaccinated for COVID-19? Fully vaccinated means getting EITHER two shots of the Pfi... = Yes”]
- Within the past two weeks
- Within in the past two months
- Within the past six months
- More than six months ago

As far as you know, are you eligible for a COVID-19 vaccine booster?
- No
- Yes
- I’m not sure.

Have you gotten a COVID-19 vaccine booster? [Display Logic: If “Are you fully vaccinated for COVID-19? Fully vaccinated means getting EITHER two shots of the Pfi... = Yes”]

No
Yes

Which vaccine did you get for your booster? [Display Logic: If “Have you gotten a COVID-19 vaccine booster? = Yes”]

Pfizer-BioNTech
Moderna
J&J-Janssen

How likely are you to get vaccinated for COVID-19 in the future? [Display Logic: If “Have you gotten at least one shot of any COVID-19 vaccine? = No”]

I definitely will NOT get vaccinated.
I probably will NOT get vaccinated.
I probably will get vaccinated.
I definitely will get vaccinated.
I am completely undecided about whether I will get vaccinated.

How likely are you to get a COVID-19 booster in the future? [Display Logic: If “Have you gotten a COVID-19 vaccine booster? = No”]

I definitely will NOT get a booster.
I probably will NOT get a booster.
I probably will get a booster.
I definitely will get a booster.
I am completely undecided about whether I will get a booster.

Have you ever tested positive for COVID-19?

No
Yes

When did you test positive for COVID-19? [Display Logic: If “Have you ever tested positive for COVID-19? = Yes”]

Within the past week
Within the past month
Within the past six months
Within the past year
More than one year ago

After testing positive, how long did you isolate at home? [Display Logic: If “Have you ever tested positive for COVID-19? = Yes”]

1-4 days
5-9 days
10-13 days
14 days or more
I did not isolate at all.

*Please answer the following two questions just to show that you're paying attention to the survey.*

Please answer “Slightly likely” to this question.

- Very unlikely
- Unlikely
- Slightly unlikely
- Slightly likely
- Likely
- Very likely

What color is the sky? Please answer this question incorrectly, on purpose, by choosing “Red” instead of “Blue.”

- Blue
- Green
- Red
- Yellow

*Please read the following text passage about guidelines for quarantine and isolation. After you read the passage, you will be asked some questions. You may take as much time as you would like to read the passage, and you may look back at the passage as you answer the questions. You may zoom in on the passage to make the text bigger.*

CDC Centers for Disease Control and Prevention

### CDC Updates and Shortens Recommended Isolation and Quarantine Period for General Population

#### Media Statement

For Immediate Release: Monday, December 27, 2021
**Contact:** Media Relations
(404) 639-3286

Given what we currently know about COVID-19 and the Omicron variant, CDC is shortening the recommended time for isolation for the public. People with COVID-19 should isolate for 5 days and if they are asymptomatic or their symptoms are resolving (without fever for 24 hours), follow that by 5 days of wearing a mask when around others to minimize the risk of infecting people they encounter. The change is motivated by science demonstrating that the majority of SARS-CoV-2 transmission occurs early in the course of illness, generally in the 1-2 days prior to onset of symptoms and the 2-3 days after.

Additionally, CDC is updating the recommended quarantine period for anyone in the general public who is exposed to COVID-19. For people who are unvaccinated or are more than six months out from their second mRNA dose (or more than 2 months after the J&J vaccine) and not yet boosted, CDC now recommends quarantine for 5 days followed by strict mask use for an additional 5 days. Alternatively, if a 5-day quarantine is not feasible, it is imperative that an exposed person wear a well-fitting mask at all times when around others for 10 days after exposure. Individuals who have received their booster shot do not need to quarantine following an exposure, but should wear a mask for 10 days after the exposure. For all those exposed, best practice would also include a test for SARS-CoV-2 at day 5 after exposure. If symptoms occur, individuals should immediately quarantine until a negative test confirms symptoms are not attributable to COVID-19.

Isolation relates to behavior after a confirmed infection. Isolation for 5 days followed by wearing a well-fitting mask will minimize the risk of spreading the virus to others. Quarantine refers to the time following exposure to the virus or close contact with someone known to have COVID-19. Both updates come as the Omicron variant continues to spread throughout the U.S. and reflects the current science on when and for how long a person is maximally infectious. These recommendations do not supersede state, local, tribal, or territorial laws, rules, and regulations, nor do they apply to healthcare workers for whom CDC has updated guidance.

Data from South Africa and the United Kingdom demonstrate that vaccine effectiveness against infection for two doses of an mRNA vaccine is approximately 35%. A COVID-19 vaccine booster dose restores vaccine effectiveness against infection to 75%. COVID-19 vaccination decreases the risk of severe disease, hospitalization, and death from COVID-19. CDC strongly encourages COVID-19 vaccination for everyone 5 and older and boosters for everyone 16 and older. Vaccination is the best way to protect yourself and reduce the impact of COVID-19 on our communities.

***The following is attributable to CDC Director, Dr. Rochelle Walensky:***

*"The Omicron variant is spreading quickly and has the potential to impact all facets of our society. CDC's updated recommendations for isolation and quarantine balance what we know about the spread of the virus and the protection provided by vaccination and booster doses. These updates ensure people can safely continue their daily lives. Prevention is our best option: get vaccinated, get boosted, wear a mask in public indoor settings in areas of substantial and high community transmission, and take a test before you gather."*

#### If You Test Positive for COVID-19 (Isolate)

| | |
|---|---|
| **Everyone, regardless of vaccination status.** | • Stay home for 5 days.<br>• If you have no symptoms or your symptoms are resolving after 5 days, you can leave your house.<br>• Continue to wear a mask around others for 5 additional days.<br><br>*If you have a fever, continue to stay home until your fever resolves.* |

#### If You Were Exposed to Someone with COVID-19 (Quarantine)

| | |
|---|---|
| **If you:**<br><br>Have been boosted<br>**OR**<br>Completed the primary series of Pfizer or Moderna vaccine within the last 6 months<br>**OR**<br>Completed the primary series of J&J vaccine within the last 2 months | • Wear a mask around others for 10 days.<br>• Test on day 5, if possible.<br><br>*If you develop symptoms get a test and stay home.* |
| **If you:**<br><br>Completed the primary series of Pfizer or Moderna vaccine over 6 months ago and are not boosted<br>**OR**<br>Completed the primary series of J&J over 2 months ago and are not boosted<br>**OR**<br>Are unvaccinated | • Stay home for 5 days. After that continue to wear a mask around others for 5 additional days.<br>• If you can't quarantine you must wear a mask for 10 days.<br>• Test on day 5 if possible.<br><br>*If you develop symptoms get a test and stay home* |

###

U.S. DEPARTMENT OF HEALTH AND HUMAN SERVICES

*CDC works 24/7 protecting America's health, safety and security. Whether disease start at home or abroad, are curable or preventable, chronic or acute, or from human activity or deliberate attack, CDC responds to America's most pressing health threats. CDC is headquartered in Atlanta and has experts located throughout the United States and the world.*

Page last reviewed: December 29, 2021

*After thinking about the information you just read in the passage above, please indicate how strongly you agree with each of the following statements.*

I trust the CDC's guidance about the COVID-19 pandemic.

- Strongly disagree
- Disagree
- Slightly disagree
- Slightly agree
- Agree
- Strongly agree

I think that the CDC's guidance is based on the best scientific evidence available.

- Strongly disagree
- Disagree
- Slightly disagree
- Slightly agree
- Agree
- Strongly agree

I think that the CDC's guidance is influenced by economic factors, such as keeping businesses up and running.

- Strongly disagree
- Disagree
- Slightly disagree
- Slightly agree
- Agree
- Strongly agree

Now imagine that we had asked you these three questions <u>BEFORE</u> the change in isolation and quarantine guidelines was announced. How would you have answered the first question, which is "I trust the CDC's guidance about the COVID-19 pandemic."

- Strongly disagree
- Disagree
- Slightly disagree
- Slightly agree
- Agree
- Strongly agree

Now imagine that we had asked you these three questions <u>BEFORE</u> the change in isolation and quarantine guidelines was announced. How would you have answered the second question, which is "I think that the CDC's guidance is based on the best scientific evidence available."

- Strongly disagree
- Disagree
- Slightly disagree
- Slightly agree
- Agree

Strongly agree

Now imagine that we had asked you these three questions BEFORE the change in isolation and quarantine guidelines was announced. How would you have answered the third question, which is “I think that the CDC’s guidance is influenced by economic factors, such as keeping businesses up and running.”

Strongly disagree
Disagree
Slightly disagree
Slightly agree
Agree
Strongly agree

*Please indicate how strongly you agree with each of the following statements about the passage.*

I think that the information in the passage is accurate and should be trusted.

Strongly disagree
Disagree
Slightly disagree
Slightly agree
Agree
Strongly agree

I think that the information in the passage is based on high-quality evidence.

Strongly disagree
Disagree
Slightly disagree
Slightly agree
Agree
Strongly agree

I think that the writing in the passage is clear and easy to read.

Strongly disagree
Disagree
Slightly disagree
Slightly agree
Agree
Strongly agree

After reading the passage, I understand what to do if I test positive for COVID-19.

Strongly disagree
Disagree
Slightly disagree
Slightly agree
Agree
Strongly agree

After reading the passage, I understand what to do if I am exposed to someone who has COVID-19.

Strongly disagree
Disagree
Slightly disagree
Slightly agree
Agree
Strongly agree

*Please answer the following questions based ONLY on the facts presented in the question and the information in the passage. You should answer based ONLY on the guidelines in the passage, even if you disagree with some of them. As a reminder, you may take as much time as you would like to read the passage, and you may look back at the passage as you answer the questions.*

James got two shots of the Pfizer-BioNTech vaccine five months ago and recently tested positive for COVID-19. After isolating at home for five days, he has a mild fever and a sore throat. What should he do?

Isolate at home for five more days, then continue his normal routine.
Isolate at home until his symptoms are gone.
Wear a mask around other people for five days, then continue his normal routine.
Wear a mask around other people until his symptoms are gone.

Patricia is unvaccinated and recently tested positive for COVID-19. After isolating at home for five days, she has no fever, and her cough and fatigue have gotten much better. What should she do?

Isolate at home for five more days, then continue her normal routine.
Isolate at home until her symptoms are completely gone.
Wear a mask around other people for five days, then continue her normal routine.
Wear a mask around other people until her symptoms are completely gone.

Mary got two shots of the Moderna vaccine last spring and a Moderna booster one month ago. Mary was recently exposed to someone who tested positive for COVID-19 but does not have any symptoms of COVID-19. Assuming Mary remains symptom-free, what should she do?

Quarantine at home for five days, then continue her normal routine.
Quarantine at home for ten days.
Quarantine at home for five days, then wear a mask around other people for five days.
Wear a mask around other people for ten days.

Casey got the J&J-Janssen vaccine four months ago and has not had a booster shot. Casey was recently exposed to someone who tested positive for COVID-19 but does not have any symptoms of COVID-19. Assuming Casey remains symptom-free, what should they do?

Quarantine at home for five days, then continue their normal routine.
Quarantine at home for ten days.
Quarantine at home for five days, then wear a mask around other people for five days.
Wear a mask around other people for ten days.

*Please answer the following questions based on YOUR OWN VACCINATION STATUS and the information in the passage. You should answer based ONLY on the guidelines in the passage, even if you disagree with some of them. As a reminder, you may take as much time as you would like to read the passage, and you may look back at the passage as you answer the questions.*

Imagine you test positive for COVID-19 on Monday. On Thursday (three days after your positive test), you have no symptoms of COVID-19. *According to the passage*, what should you do that day?

Isolate at home and avoid other people.
Wear a mask when around other people.
Wear a mask only when attending large indoor gatherings.
Make no changes to your normal routine.

Imagine you test positive for COVID-19 on Monday. On the following Monday (seven days after your positive test), you have a mild fever and a dry cough. *According to the passage*, what should you do that day?

Isolate at home and avoid other people.
Wear a mask when around other people.
Wear a mask only when attending large indoor gatherings.
Make no changes to your normal routine.

Imagine you are exposed to someone who has COVID-19 on Monday. On Thursday (three days after exposure), you have no symptoms of COVID-19. *According to the passage*, what should you do that day?

Quarantine at home and avoid other people.
Wear a mask when around other people.
Wear a mask only when attending large indoor gatherings.
Make no changes to your normal routine.

Imagine you are exposed to someone who has COVID-19 on Monday. On the following Monday (seven days after exposure), you have no symptoms of COVID-19. *According to the passage*, what should you do that day?

Quarantine at home and avoid other people.
Wear a mask when around other people.
Wear a mask only when attending large indoor gatherings.
Make no changes to your normal routine.

*Now please answer the same four questions based on what YOU PERSONALLY think you would do in each situation (even if that is different from what the passage says you should do).*

Imagine you test positive for COVID-19 on Monday. On Thursday (three days after your positive test), you have no symptoms of COVID-19. What *would* you do that day?

Isolate at home and avoid other people.
Wear a mask when around other people.
Wear a mask only when attending large indoor gatherings.

Make no changes to your normal routine.

Imagine you test positive for COVID-19 on Monday. On the following Monday (seven days after your positive test), you have a mild fever and a dry cough. What *would* you do that day?

Isolate at home and avoid other people.
Wear a mask when around other people.
Wear a mask only when attending large indoor gatherings.
Make no changes to your normal routine.

Imagine you are exposed to someone who has COVID-19 on Monday. On Thursday (three days after exposure), you have no symptoms of COVID-19. What *would* you do that day?

Quarantine at home and avoid other people.
Wear a mask when around other people.
Wear a mask only when attending large indoor gatherings.
Make no changes to your normal routine.

Imagine you are exposed to someone who has COVID-19 on Monday. On the following Monday (seven days after exposure), you have no symptoms of COVID-19. What *would* you do that day?

Quarantine at home and avoid other people.
Wear a mask when around other people.
Wear a mask only when attending large indoor gatherings.
Make no changes to your normal routine.

*Please answer the following questions with your <u>BEST GUESS</u>.*

*As a reminder, effectiveness of a vaccine can be defined based on three different factors:*

*1) how good it is at preventing* infection *with COVID-19,*
*2) how good it is at preventing* hospitalization *from COVID-19, and*
*3) how good it is at preventing* death *from COVID-19.*

*Each of these effectiveness numbers can be different.*

As you read in the passage, vaccination "decreases the risk of severe disease, hospitalization, and death from COVID-19." If you had to guess, how effective **right now** is a COVID-19 vaccine <u>WITHOUT</u> a booster at preventing *hospitalization* from COVID-19?

90% or more
80-89%
70-79%
60-69%
50-59%
40-49%
30-39%
20-29%
10-19%
Less than 10%

As you read in the passage, vaccination "decreases the risk of severe disease, hospitalization, and death from COVID-19." If you had to guess, how effective **right now** is a COVID-19 vaccine WITHOUT a booster at preventing *death* from COVID-19?

90% or more
80-89%
70-79%
60-69%
50-59%
40-49%
30-39%
20-29%
10-19%
Less than 10%

As you read in the passage, vaccination "decreases the risk of severe disease, hospitalization, and death from COVID-19." If you had to guess, how effective **right now** is a COVID-19 vaccine WITH a booster at preventing *hospitalization* from COVID-19?

90% or more
80-89%
70-79%
60-69%
50-59%
40-49%
30-39%
20-29%
10-19%
Less than 10%

As you read in the passage, vaccination "decreases the risk of severe disease, hospitalization, and death from COVID-19." If you had to guess, how effective **right now** is a COVID-19 vaccine WITH a booster at preventing *death* from COVID-19?

90% or more
80-89%
70-79%
60-69%
50-59%
40-49%
30-39%
20-29%
10-19%
Less than 10%

If you had to guess, do you think the guidelines for isolation and quarantine will be changed again in the next six months?

Definitely not

Probably not
Probably
Definitely
I'm not sure.

The passage says “that vaccine effectiveness against infection for two doses of an mRNA vaccine is approximately 35%.” As more data about the Omicron variant becomes available, do you think that number is likely to change?

Definitely not
Probably not
Probably
Definitely
I'm not sure.

The passage says, “A COVID-19 vaccine booster dose restores vaccine effectiveness against infection to 75%.” As more data about the Omicron variant becomes available, do you think that number is likely to change?

Definitely not
Probably not
Probably
Definitely
I'm not sure.

*Please answer the following questions about your background.*

How old are you?

18-24
25-29
30-39
40-49
50-64
65 or older

Which best describes your gender? Please choose as many options as apply.

Female
Male
Non-binary
Transgender
Another option not listed here (please specify): _________

Are you Hispanic or Latino/Latina/Latinx?

Not Hispanic or Latino/Latina/Latinx
Hispanic or Latino/Latina/Latinx

Which best describes your race? Please choose as many options as apply.

American Indian or Alaska Native

Asian
Black or African American
Native Hawaiian or Other Pacific Islander
White
Another option not listed here (please specify): _________

What is your *highest* level of formal education?
I do not have a high school diploma.
I have a high school diploma (or equivalent).
I went to college but don't have a degree.
I have an associate's degree
I have a bachelor's degree.
I have a graduate or professional degree.

Generally speaking, which of the following best describes your political affiliation?
Strong Republican
Leaning Republican
Leaning Democratic
Strong Democratic
Independent
Another option not listed here (please specify): _________

In which state do you live?
▼ Alabama ... District of Columbia

How would describe the area where you live?
Rural area
Suburban area
Urban area

Did you use Google or any other outside sources to answer the questions? Please answer honestly. Your payment does NOT depend on your response to this question.
No
Yes

How carefully did you complete this survey? Please answer honestly. Your payment does NOT depend on your response to this question.
Not at all carefully
Slightly carefully
Moderately carefully
Carefully
Very carefully
